# The Relationship between Democracy embracement and COVID-19 reported casualties worldwide

**DOI:** 10.1101/2021.01.11.21249549

**Authors:** Muhammad R. Hussein, Thamer AlSulaiman, Mohamed Habib, Engy A. Awad, Islam Morsi, John R. Herbold

**Affiliations:** Uiversity of Texas UTHealth School of Public Health; University of Iowa Department of Computer Science; Science Collaboration Development Center; German University in Cairo; University of Texas UTHealth School of Public Health

## Abstract

**Background:** The COVID-19 toll of cases and deaths followed an uneven pattern across the world. The literature has partly explained the observed discrepancy between the different countries by country-specific and systemic patterns worldwide. In this study, we propose an additional explanation that the magnitude of COVID-19 toll reported to the WHO could be influenced by the level of free speech and Democracy in the reporting countries.

**Methods:** We constructed a longitudinal dataset including the daily COVID-19 count of cases and deaths worldwide and each country’s respective score on the Freedom in the World index. We applied two Generalized Estimating Equation models to investigate if a country’s reported toll count of COVID-19 cases and deaths is related to that country’s freedom level. We controlled for factors identified in the current literature to affect the pandemic’s spread.

**Results:** A country’s score on the Freedom In the World Index was associated with its reported COVID-19 cases count (57028.43, 95% CI 985.3619 - 113071.5, P= 0.0461) and deaths count (3473.273, 95% CI1217.12-5729.42, P=.002). Also, despite having almost equal shares of the world’s population, countries at the bottom category of the Freedom index reported 21% and 11% of the COVID-19 toll cases and death counts reported by countries of highest scores on the index, respectively.

**Conclusions:** The known magnitude of the COVID-19 pandemic’s morbidity and mortality appears to be as transparent as the reporting countries uphold free speech and Democracy. This pattern could potentially misguide international aid and global vaccine distribution plans.

## Introduction

Concurrent to the news on COVID-19 vaccines, the question has arisen on the order of priority in distributing them. On the national level in the United States, the CDC was prompt to establish broad vaccine distribution phases.^1^ On the international level, the debate is escalating on balance between some countries’ financial ability to secure their vaccine supplies and the needs of mid and low-income countries.^2^ A critical, yet not fully verified, assumption basing many of the involved public health and logistic considerations is that the information we have on the COVID19 global spread is independent and transparent enough to be an accurate representation of its real impact. In this research, we examined the validity of this assumption on the global level.

Since the beginning of the epidemic, multiple factors cast doubt on the COVID-19 toll calculations’ accuracy in different countries. There was a lack of consistency in selecting high-risk population groups that some countries initially monitored for COVID-19.^3^ Government authorities worldwide used various combinations of clinical, epidemiological, and laboratory confirmation requirements to determine which cases constitute COVID-19 cases as the primary ailment.^4^ The types, availability, and accuracy of the testing kits used in screening for COVID-19 varied worldwide at different time points. Eventually, most countries used various diagnostic tools of different sensitivities though not all countries could always have enough testing kits. Manufacturing limitations sometimes added to the uncertainty on some of the available kits.^5^

Despite these considerable differences between the individual countries, there have been observable systematic patterns in the COVID-19 impact numbers on the national and international levels, revealing socio-economic challenges for minorities and disadvantaged populations. For example, in the US, minority populations were disproportionately impacted by the COVID-19 epidemic, prompting questions on the health equity considerations and barriers to care.^6^ At the global level, other systemic patterns appeared in the COVID-19 toll of cases and deaths, such as the seemingly paradoxical observation that the epidemic could be spreading at higher rates in developed and prosperous countries than others of weaker health care infrastructure.^7^

Another observation, which the current literature has not fully explained yet, is the discrepancy between countries in their counts of COVID-19 cases and deaths.^8^ The full explanation of this observation could reveal what makes a country more vulnerable to COVID-19 and which countries are the hotspots of the epidemic and logistic priorities in vaccine distribution.^9^ The literature has proposed factors of weather, location, health care infrastructure, aging population, and others. These explanations could only partly decipher the epidemic’s spread pattern. ^10^ In this study, we proposed an additional explanation that identifies a potential barrier to equitable channeling of support, including vaccines, to genuine need populations. Our research hypothesis is that the magnitude of a country’s reported toll of COVID-19 cases and deaths could be under the influence of the degrees of Freedom of speech and Democracy that country embraces. The less a government upholds these governance elements, the lower impact it could be reporting about the epidemic in its people. The premises of this hypothesis are the inconsistent spread of COVID-19 across the globe without a full explanation, multiple media reports on governments considering independent reporting on COVID-19 as misinformation, and studies from independent research institutions highlighting a step up in governmental control and a decline in Democracy in around the world during the pandemic. ^11,12,13,14^

On top of misguiding scientific efforts, this possibility would likely create a barrier to equitable access to the COVID-19 vaccines. The perceived low impact of the epidemic in some countries could mislead the distribution efforts championed by the international organizations and the vaccine manufacturers into deprioritizing sending vaccine aid to these populations, contrary to their actual need.

To investigate this hypothesis, we conducted this longitudinal analysis of the potential relationship between the toll of cases and deaths reported by the world countries and their respective scores on the Freedom In the World Index, recognized in the politico-economic literature as a measure of Democracy and Free speech around the globe.

## Study Data And Methods

The observation unit was daily country. We collected records of confirmed COVID-19 impact reported to the WHO from world countries. Our study dataset tracked 182 countries over 242 days from January 23, 2020, to September 20, 2020.

### Outcome of Interest

We examined the cumulative daily counts of confirmed COVID-19 cases and deaths from the COVID-19 dashboard provided by the Center for Systems Science and Engineering (CSSE) at Johns Hopkins University. This online repository hosts data on COVID-19 deaths and cases worldwide, collected from official and independent sources.

### Predictor variable

We used Freedom In the World Index (2020) to represent Democracy and Free speech in the world countries^13^. This annual index is developed by the US-based independent research institution Freedom House (US). The index is a composite score on a scale from one to a hundred, aggregating each country’s sub-scores on a wide array of surveyed measures of Democracy such as political pluralism, Freedom of expression, the rule of law, and individual rights. Each country is also designated a Freedom In the World status (Free, Partly Free, or Not Free) based on its respective aggregate scores on Political Rights and Civil Liberties.^15^

### Covariates

We added control variables representing the different factors hypothesized in the current literature to influence the spread of COVID-19. The model included fixed effects for daily temperature and ultraviolet index levels, sourced from WeatherStack data repository, which collects worldwide weather data from stations worldwide.^16^ Additionally, we added fixed effects of worldwide enactment of containment measures and public health policies, represented by daily records in Government Stringency Index developed by Oxford University s.^17^

We also added fixed effects for pollution levels (in mean annual exposure in micrograms per cubic meter, 2017),^18^ population density (2019), and wealth (in the 2019 gross domestic product in PPP). We obtained this information from the World Bank Databank. Finally, we used the Health Access and Quality Index (2016), which scores the accessibility of health care services in the different countries, as a proxy of the accessibility to care during the COVID-19 epidemic.

## Methods

We estimated the relationship between the countries’ scores on the Freedom In the World index and their reported cumulative count of COVID-19 cases and deaths, using a Generalized Estimating Equation (GEE) model with an independent correlation structure. The model included the aforementioned covariates believed, in the consensus of literature in multiple scientific disciplines, to influence the global spread of COVID-19. We used SAS 9.4 TS Level 1 M6 to conduct the statistical analysis and impute the missing data. Although multiple imputations is not a required step for GEE models, for being population-based models, the consensus in Biostatistics literature is that imputing the missing data improves the adequacy of the GEE parameter estimates.

## Results

### Descriptive statistics

In the Free category, the highest scores on the Freedom In the World Index, there are 72 countries. This category collectively hosts 38.28% of the world population and reported 72.37% of the world’s COVID-19 caseload and 78.77% of the world’s COVID-19 related deaths. Countries in this category produced 56.56% of the world’s 2019 GDP in PPP and had an average score of 76.94 on the Health Access and Quality index. (Figure 1)

**Figure 1.**
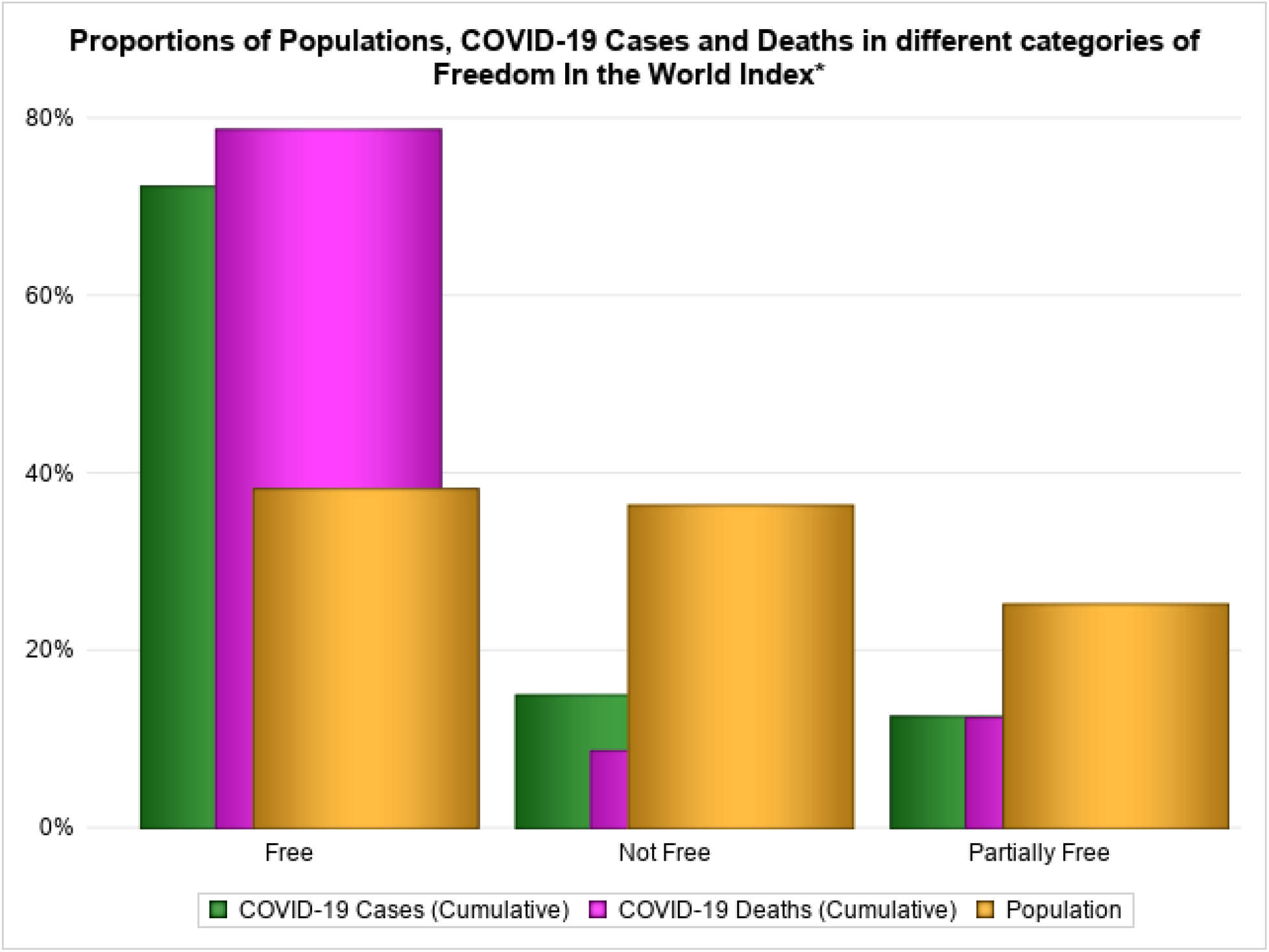

In the Not Free category, the lowest scores category, there are 48 countries. They collectively host 36.3% of the world population and accounted for 15% of the total world COVID-19 cases and 8.7% of the total deaths. These countries produced 29.66% of the world GDP in PPP and had an average score of 51.81 on the Health Access and Quality index. (Figure 1)

### Inferential statistics

There was a statistically significant positive association between a country’s score on the Freedom In the World Index and the cumulative count of cases that the government reported. For every point decrease on the index, the average country reported 57028 less cumulative COVID-19 cases, and vice versa (57028.43, 95% CI 985.3619 - 113071.5, P= 0.0461). There was also a statistically significant positive association between a country’s score on the index and the cumulative count of deaths it reported. For every point decrease on the index, the respective country reported 3473.27 less COVID-19 deaths, cumulative, over the study period (3473.273, 95% CI1217.12-5729.42, P=.002). (Figure 2)

**Figure 2.**
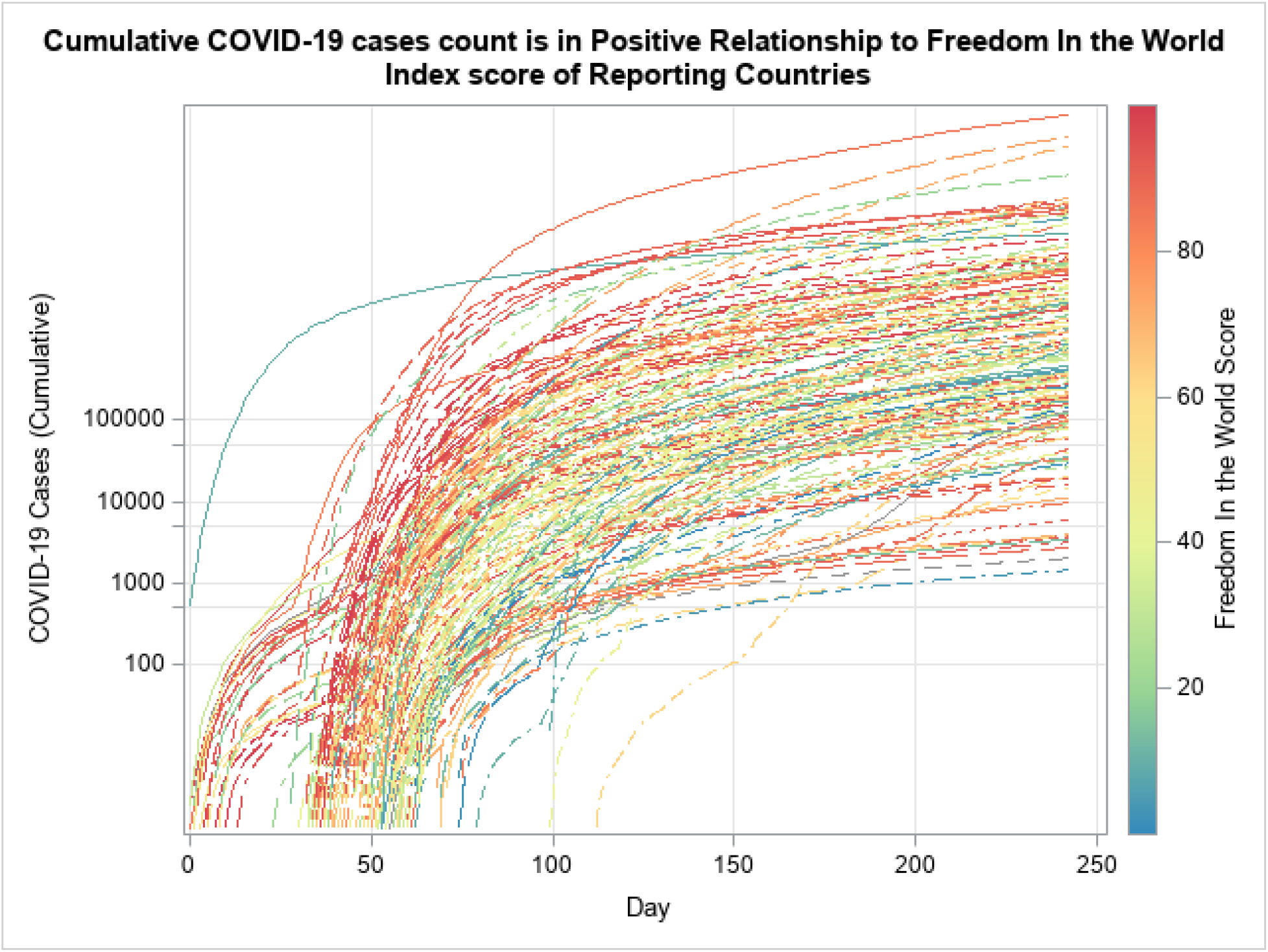

## Discussion

The results suggest a relationship between the COVID-19 toll reported to the World Health Organization from the different countries and the respective countries’ levels of Freedom. The less a country embraced Democracy and Free speech, the lower the magnitude of the COVID-19 morbidity and mortality it reported to the world about its people. Countries in the Not Free category reported shares of global COVID-19 cases and deaths were 21% and 11% of the counts reported by the Free category, respectively, despite that the two categories have almost equal shares of the world population. Paradoxically, the Not Free category had poorer economy and weaker health care infrastructure, in terms of access and quality, than the Free category. This discrepancy is independent of the factors believed to influence the pandemic’s spread, such as access to health care, containment measures and public health policies, weather conditions, pollution, demographic characteristics, or wealth.

These findings cast doubt on the validity of the assumption that the observed toll of COVID-19 morbidity and mortality counts is transparent enough to represent the reality of the epidemic’s impact accurately. Without a valid representation of the COVID-19 reality, scientific efforts could investigate inaccurate findings and drive unreliable conclusions, leading to a misallocation of global aid resources.

For example, the currently available information led some research efforts to investigate a perceived observation that the pandemic has not affected low-income countries as it did with high-income countries. One explanation was that rich countries could afford broader screening efforts, which will ultimately detect a higher number of cases than other countries. Yet, this study’s findings could propose an entirely different explanation considering that many low-income countries also have low embracement of Democracy.

Another example is the apparent association between colder temperatures and the spread of the virus, which led some to believe that the summer would restrain the epidemic early in the epidemic’s timeline.^19^ Some researchers suggest that this misperception probably deterred the COVID-19 response in the summer of 2020 by encouraging complacency in following social distancing measures, at least in part.^20^ Yet, in the light of the findings in this study, we could consider the possibility that the low COVID-19 impact in hot countries in areas like Africa could be partially coming from a lack of Freedom of speech and low Democracy.

In future consideration, the perceived impact of the epidemic could be a key element in allocating international aid and vaccine quotas to developing countries. Considering the findings, the international organizations and vaccine manufacturers will likely deprioritize aid to some countries based on their low toll of COVID-19 cases and deaths, which could be, at least in part, because of a lack of transparency in reporting the reality of the COVID-19 situation in them. This role of lack of Democracy as a barrier against access to care could be a new socio-economic determinant of health care in the COVID-19 epidemic. Further research could help bridge this barrier by considering various cultural and socio-economic factors that could influence a country’s government ability and willingness to help the international community get a realistic understanding of the epidemic. While such scientific inquiry goes beyond this study’s scope, this study’s findings could be a useful starting point for decision-makers and members of the research community working to promote equitable access to the global pool of therapeutic and preventive aid resources.

## Limitations

This study has several limitations. First, the inconsistencies between the standards that different countries have followed to varying points of time in the study period could introduce inaccuracies in the COVID-19 toll count of cases and deaths used in the analysis. Second, we did not obtain information on the availability of COVID-19 testing in different countries, which might shape the observed spread pattern. Third, the weather information used in the analysis is on each country’s political capital, which might not be fully representative of the weather in all that country’s regions. Fourth, the study’s analysis does not divulge the causality considerations underlying the findings, which could be an area for further research.

## Data Availability

The authors will gladly made the study data available by request.

## Declaration of Conflicting Interests

The authors declared no potential conflicts of interest with respect to the research, authorship, and/or publication of this article.

## Notes

### Competing Interest Statement

The authors have declared no competing interest.

### Funding Statement

No external funding was received

### Author Declarations

No IRB approval was necessary.

